# Phenotyping aldehyde metabolism in humans uncovers novel genetic mutations in aldehyde dehydrogenase 2

**DOI:** 10.1101/2020.06.26.20137513

**Authors:** Freeborn Rwere, Joseph R. White, Xiaocong Zeng, Leslie McNeil, Kevin N. Zhou, Martin S. Angst, Che-Hong Chen, Daria Mochly-Rosen, Eric R. Gross

## Abstract

Aldehydes are toxic chemicals rapidly metabolized by the enzyme aldehyde dehydrogenase 2 (ALDH2). Nearly 8% of the world population carry an ALDH2 genetic variant *rs671* (E504K) causing inefficient aldehyde metabolism recognized by facial flushing after alcohol consumption. Importantly, *rs671* carriers have a several-fold increased risk of cancer, neurodegenerative, and heart disease attributed to inefficient aldehyde metabolism. Although *rs671* is common, it is unlikely *rs671* is the only inactivating ALDH2 mutation in humans.

By recruiting human volunteers reporting facial flushing after alcohol consumption, we identified previously uncharacterized ALDH2 point mutations including *rs747096195* (R101G), *rs190764869* (R114W), and *rs4646777* (G>A). Using ALDH2 recombinant enzyme, we determined R101G and R114W have a reduced efficiency in acetaldehyde metabolism unique to E504K (V_max_/K_m_, 1.95±0.01^+^ and 2.77±0.02^+^ relative to 0.93±0.03, respectively n=3/group, ^+^P<0.001 relative to 7.97±0.51 for wild type ALDH2).

When subjecting human volunteers to an alcohol challenge (0.25g/kg), acetaldehyde levels increased 9-fold for *rs671* (E504K), 2-fold for *rs747096195* (R101G) and *rs190764869* (R114W) and was no different for *rs4646777* (G>A) relative to volunteers carrying wild type ALDH2. Further, for volunteers tested, heart rate changes after alcohol strongly correlated with decreased acetaldehyde metabolism (r=0.90, n=26) as opposed to facial flushing. Taken together, we identified novel inactivating ALDH2 point mutations and developed a non-invasive diagnostic test to phenotype aldehyde metabolism in humans. Identifying ALDH2 genetic differences and their phenotype in response to alcohol can provide more precise individual recommendations regarding the health risks of aldehyde exposure.

## Introduction

Aldehydes are toxic compounds that can be inhaled, consumed, or endogenously produced^1-4^. Exogenous exposures of aldehydes, such as acetaldehyde and formaldehyde, can occur from sources such as air pollution, cigarette smoke, and alcohol^2^. Endogenous sources of aldehydes, such as 4-hydroxynonenal and malondialdehyde, are produced from lipid peroxidation such as during surgery or organ injury^3, 4^. As aldehydes are highly diffusible and penetrate membranes, aldehydes intracellularly can cause aldehyde-induced DNA and protein adducts that are mutagenic, carcinogenic, and cytotoxic which in turn drive the pathogenesis for cancer, cardiovascular, and neurodegenerative diseases^5, 6^.

A major protective mechanism the cell has against aldehyde-induced toxicity is the enzyme aldehyde dehydrogenase 2 (ALDH2) which acts to neutralize and limit aldehyde-induced damage^3^. However, nearly 540 million people world-wide, mainly of East Asian descent, carry an ALDH2 genetic variant *rs671* (E504K) decreasing ALDH2 enzymatic activity by at least 50% relative to the wild type ALDH2 enzyme^7^. With aldehyde exposures, *rs671* is less efficient in metabolizing aldehydes. Subsequently, aldehyde accumulation and intracellular aldehyde-induced adduct formation occurs in greater frequency relative to those carrying the wild type ALDH2 enzyme; leading to higher risks of cancer, cardiovascular, and neurodegenerative diseases for people carrying *rs671*^4, 8-10^.

Although *rs671* is common, it is unlikely the only inactivating ALDH2 mutation. In humans, several additional ALDH2 missense mutations, including a common intron mutation (*rs4646777*, G>A, 0.18 frequency), are identified by whole genome sequencing^11^. However, whether these additional ALDH2 mutations are inactivating mutations are unknown. As such, screening ALDH2 mutations for those causing inefficient aldehyde metabolism is important considering the health risks associated with the *rs671* variant and aldehyde exposure^12^.

Facial flushing after alcohol can be an indicator of aldehyde accumulation. For those carrying *rs671*, the acetaldehyde accumulation after alcohol consumption produces facial flushing and tachycardia^8^; a condition called Alcohol Flush Syndrome^13^. However, it is also unlikely that all people who have facial flushing after alcohol accumulate acetaldehyde. Thus, a means to identify people who accumulate aldehydes due to carrying an ALDH2 point mutation is important considering the health risks associated with *rs671*. Here we recruited human volunteers self-reporting facial flushing after alcohol consumption, characterized ALDH2 mutations identified using recombinant protein, and developed a non-invasive screening method to phenotype aldehyde metabolism in humans through using breath monitoring and physiological measurements.

## Methods

Prior to recruitment and testing, IRB approval was obtained from Stanford University (IRB 46095). During IRB review, this study was identified as a basic experimental study involving humans with the manipulation or task used (consuming alcohol) expressly used for measurement and was not considered an intervention. Written informed consent was received from participants prior to inclusion in the study. The experiments conformed to the principles set out in the WMA Declaration of Helsinki and the Department of Health and Human Services Belmont Report.

### Participant recruitment

Participants were recruited from December 2018 through March 2020 using flyers posted on the Stanford University campus in succession. The initial flyer recruited people who flush after they consume alcohol for an alcohol study. During the recruitment phase, public support for this project triggered the recruitment flyer at Stanford University to be posted on the Facebook group page Subtle Asian Traits that has ∼1.8 million members. Due to the public response caused by the flyer posted on a Facebook group page, the flyer was modified to recruit additional people only of non-Asian descent.

Participants were initially screened for exclusion using a questionnaire (see Supplemental Material which describes exclusion criteria). Participants were also instructed to complete a CAGE questionnaire to screen out participants with potential alcoholism. Those participants eligible then provided a saliva sample for DNA extraction and purification (Quick-DNA Miniprep Kit, Zymo Research). DNA was amplified using 12 primers designed to cover the ALDH2 exome for sequencing while in addition an ALDH2 common intron mutation (*rs4646777*) as described (Supplemental Table 1). After amplification, samples were sent for Sanger sequencing (Eton Bioscience).

### *In Silico* ALDH2 Modeling

Protein data bank (PDB) entry 1O02 was imported into PyMOL (Schrödinger, Inc.) for molecular visualization of wild-type ALDH2 and to mutate R101 and R114 by using the protein mutagenesis function.

### ALDH2 Site-Directed Mutant Construction, Protein Expression and Purification

Based upon genotyping results, site-directed mutagenesis of human ALDH2 was performed using a QuikChange site-directed mutagenesis kit (Agilent Technologies) using the kanamycin resistance template plasmid, pET-28a-c (+). Oligonucleotides used to generate ALDH2 mutants (R101G, R114W and E504K) were purchased (Integrated DNA Technologies). Co-expression of wild-type ALDH2 and its site directed mutants (R101G, R114W and E504K) was achieved using the pEDuet-1 vector plasmid (Millipore Sigma, Milwaukee, WI). One copy of wild-type ALDH2 was cloned into the His-tag cloning site of the pETDuet-1 vector plasmid by EcoRI and HindIII, whereas another copy was cloned into the S-tag using NdeI and XhoI. For co-expression of wild-type ALDH2 with the variants, one copy of wild type was cloned into the His-tag cloning site as described above, whereas the variants were cloned into the S-tag cloning site of pETDuet-1 plasmid using NdeI and XhoI. Co-expression of the recombinant proteins was carried out at 30 °C in the presence of an ALDH2 chaperone using *E. coli* BL21 (DE3) host cells. 0.5 M IPTG was used for induction at 30°C for 16 hours. Purification of the recombinant proteins was achieved using the His-trap nickel affinity column (GE Healthcare).

### ALDH2 Enzymatic Activity Assay

NAD^+^ conversion to NADH in the presence of acetaldehyde was used to determine ALDH2 enzyme activity as previously described ^14^. A blank control was run without acetaldehyde. Results were collected in triplicates on two separate days.

### Human Alcohol Challenge

Participants were selected for the alcohol challenge portion of the study based on genotyping results. At this stage, participants were also excluded if homozygous for *rs671* (secondary to the expected severity of the alcohol response). Participants heterozygous for *rs671* (E504K) were called back for participation in the alcohol challenge in addition to those without sequenced mutations were selected to closely match the age, sex and weight of the participants heterozygous for *rs671* (E504K). Additional participants were selected for the alcohol challenge study based upon genotyping results identifying point mutations including *rs747096195* (R101G), *rs190764869* (R114W), and *rs4646777* (G>A, intron 8).

Participants were instructed prior to the alcohol challenge to not eat or drink for 2 hours prior to testing. Testing was performed between 12pm and 4pm on a weekday. Upon arrival, patients were consented and weighed. To assess physiologic measurements, a five lead EKG was placed and EKG measurements were recorded (ADInstruments). Blood pressure and heart rate was measured using a standard blood pressure cuff and pulse oximeter (DynaMap). Skin temperature was measured under the left cheek bone using liquid crystal technology (Crystaline II Temperature Indicators, Sharn Anesthesia). Baseline hemodynamic data including blood pressure, heart rate, and skin temperature were collected. Prior to testing, a hand-held alcohol breath test meter confirmed no alcohol was consumed before testing (BACtrack Element Pro). Breath samples were collected by a 3L Tedlar bag (Zefon International) that were sealed after collection. Breath samples were analyzed within 4 hours of sample collection.

After establishing a baseline, participants were given a 415 mL (14oz) drink consisting of 0.25g/kg Ketel One vodka, water, and Minute Maid lemonade. For reference in the United States, a standard drink is 14 grams of alcohol which is equal to one 12 oz glass of 5% beer or one 5oz glass of 12% wine ^15^. As an example, a 70kg person in this study drank 17.5g of alcohol. Participants consumed the first 7oz of the 14oz drink during the first 5 minutes of the study and the second 7oz during the second 5 minutes. After baseline measurements taken 3 minutes before alcohol consumption, physiologic measurements of heart rate, blood pressure, skin temperature, and breath metabolites were taken at 5, 10, 15, 30, 60, 90, 120, and 150 minutes during and after alcohol consumption (Figure 2A). After completion of the study, a breath alcohol test and a standard field sobriety test was administered. The study lasted a total duration of 3 hours. When collecting the data, we considered the heart rate, blood pressure and facial skin temperature as the physiological measurements to analyze in relation to the acetaldehyde levels measured. Participants completing the alcohol challenge were given $125 compensation.

**Figure 1.**
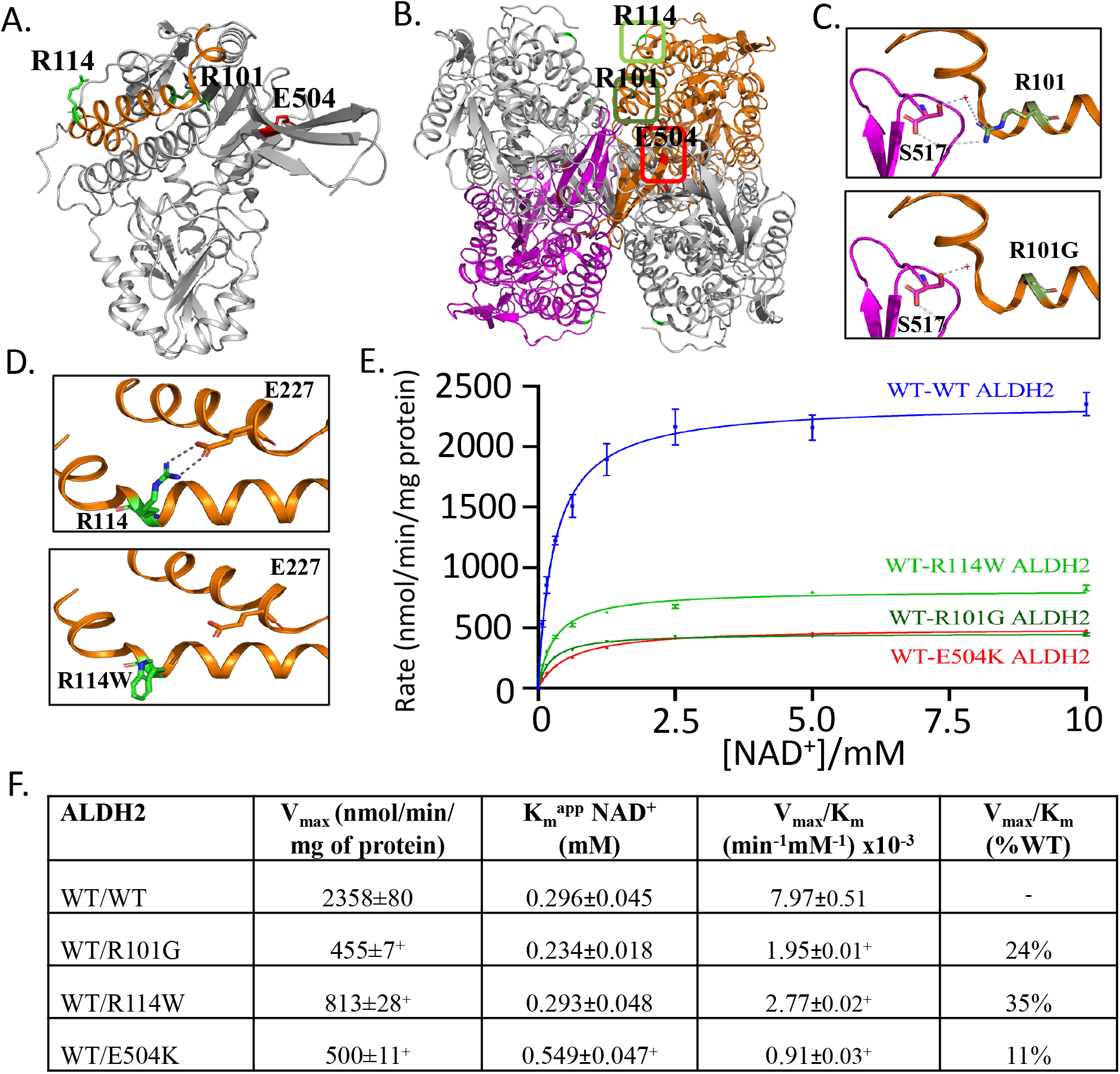
Missense mutations ALDH2 R101G and R114W cause inefficient acetaldehyde metabolism. **A**. Crystal structure of ALDH2, with 3 amino acids within the structure highlighted, R101G (*rs747096195*, dark green), R114W (*rs190764869*, light green) and E504K (*rs671*, red). **B**. ALDH2 tetramer with location of R101 (dark green box) and R114 (light green box). **C**. R101G point mutation results in loss of a hydrogen bond involved in stabilizing ALDH2 dimerization. **D**. R114W point mutation results in loss of hydrogen bonds between the adjacent alpha helix within the ALDH2 monomer. **E**. *In vitro* results supporting inefficient acetaldehyde metabolism for R101G and R114W. **F**. Summary table of V_max_ and K_m_ for missense mutations R101G and R114W relative to wild type ALDH2 and E504K. n=3/group, ^+^P<0.01 versus wild type ALDH2 enzyme

**Figure 2.**
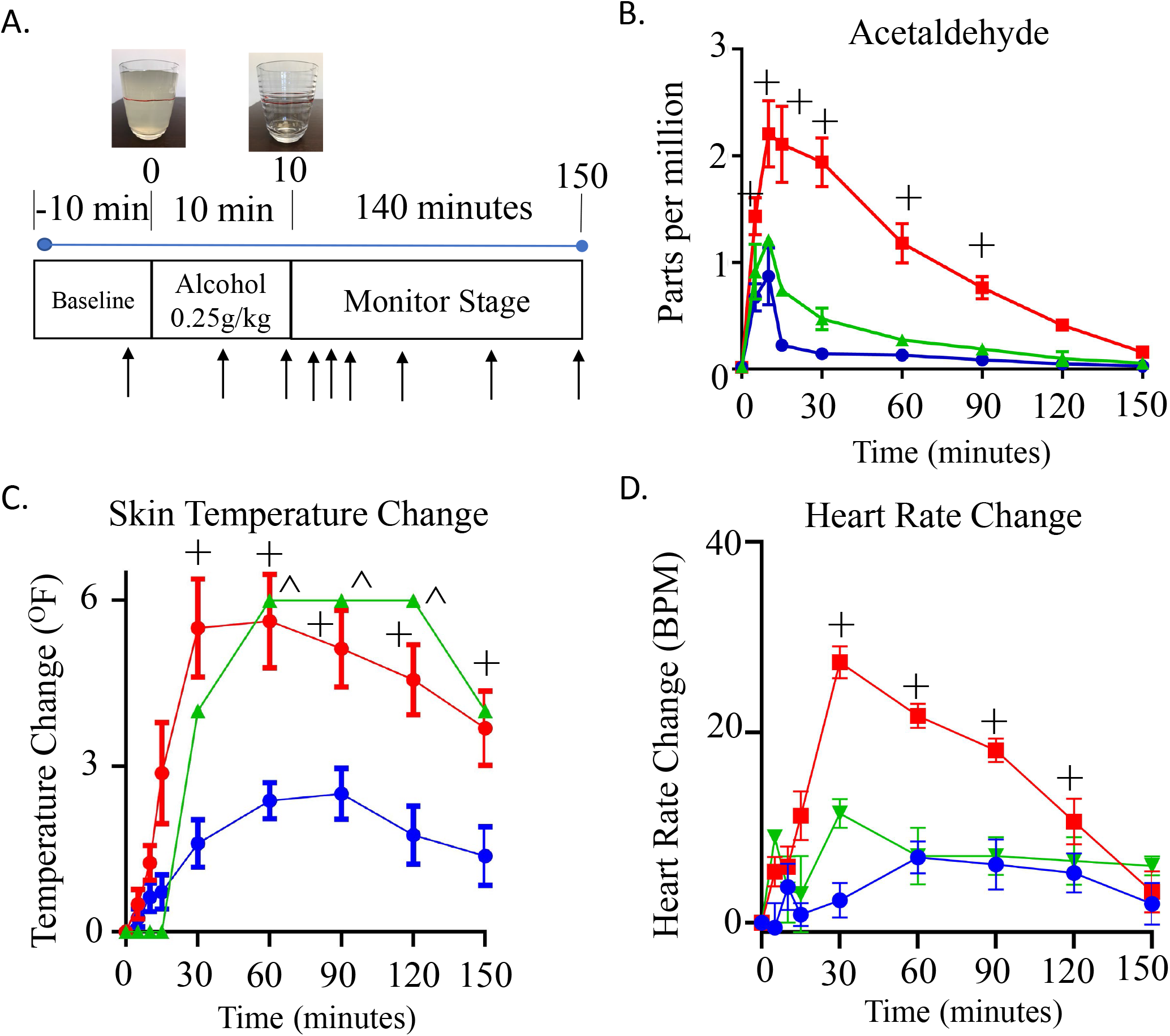
Experimental protocol, breath metabolite and physiologic characteristics after alcohol challenge. **A**. Alcohol challenge protocol. After 10 minutes baseline, subjects were given an alcohol challenge (0.25g/kg); consumed over 10 minutes. Subjects were monitored by measuring physiologic parameters. Breath measurements were measured at 9 time points (black arrow). After collecting a baseline measurement, the next 2 measurements were during alcohol consumption followed by monitoring for an additional 140 minutes after alcohol consumption. **B**. Breath concentration (in parts per million) of acetaldehyde. **C**. Change in skin temperature after alcohol consumption. **D**. Change in heart rate after alcohol consumption. For all panels, blue lines are wild type ALDH2 subjects (n=8), red lines are subjects that are heterozygotes for *rs671* (E504K, n=8), green are heterozygotes for either *rs190764869* (R101G) or *rs747096195* (R114W). ^+^ or ^^^P<0.01 versus wild type ALDH2 subjects.

### Breath metabolite analysis

The breath metabolite acetaldehyde was measured from a Tedlar bag using selective ion flow mass spectrometry within 4 hours of collection (Syft Voice Ultra, New Zealand). Soft ionization was carried out using H_3_O^+^ and NO^+^ with the mass and reaction ratios described for measuring acetaldehyde (Supplemental Table 2). Helium was used a carrier gas.

### Statistical Analysis

All data was presented as mean ± SEM. The number needed to recruit for this study was based upon a prior study examining differences in acetaldehyde metabolism in the blood. Based upon the dose of alcohol (0.25g/kg) producing a 5-fold difference in blood acetaldehyde levels, we predicted that at a minimum a need to recruit 4 people per group for statistical differences between people that are wild type ALDH2 relative to people that are heterozygous for *rs671* (E504K)^16^. For analysis of differences in metabolites or physiological data, a two-way ANOVA followed by Bonferroni correction for multiplicity was used in order to compare each group to the control group or to compare measurements performed within a group over time. When comparing heart rate changes with acetaldehyde levels, a Pearson correlation test with a 2-tailed p-value was used. Statistical analysis was performed using GraphPad Prism 6. ^+^ or ^=*p* < 0.01 was considered statistically significant between groups.

## Results

Initially, 51 people participated in a phone screen, 29 people provided a saliva sample for genotyping, and 16 people were given an alcohol challenge where 8 were heterozygotes for *rs671* and 8 were not (Supplemental Figure 1A). After the recruitment flyer was modified to recruit participants of non-Asian descent, 68 additional people underwent a phone screen, 37 people provided saliva samples, and based on the genotyping results, 10 people were given an alcohol challenge (Supplemental Figure 1B).

After genotyping, we identified people heterozygous for *rs671* (E504K), *rs747096195* (R101G), *rs190764869* (R114W) and an intron *rs4646777* (G>A). When examining the crystal structure, R101G and R114W are located in distinct regions of the α-helix that are separate from E504K (Figure 1A). A closer look at the ALDH2 tetramer suggests that R101 is important in the interaction between ALDH2 subunits while R114 is important in the interaction of alpha helices within the same subunit (Figure 1B). When R101 is mutated to a glycine, a hydrogen bond *via* a conserved water molecule is lost between R101 and S517 (Figure 1C). Upon R114W mutation, the two hydrogen bonds between E227, present within the same subunit on an adjoining alpha helix, are lost (Figure 1D). To mimic the heterozygous condition in humans, we co-expressed wild type and the ALDH2 point mutations into the pETDuet-1 vector. The co-expressed mutants R101G, R114W, and E504K had a significant decrease in V_max_ compared to wild-type ALDH2 (Figure 1E and 1F, 455±7^+^, 813±28^+^, and 500±11^+^ versus 2358±80 nmol/min/mg protein for wild type ALDH2, respectively, n=3/group). Further, the K_m_ for R101G and R114W co-expressed with wild-type ALDH2 were similar to wild type ALDH2 (Figure 1E and 1F, K_m_ = 0.234±0.018 and 0.293±0.045 versus 0.296±0.045mM, respectively). However, E504K co-expressed with wild type ALDH2 had a ∼2-fold increase of K_m_ compared to wild type, R101G and R114W ALDH2 (Figure 1F).

To explore the phenotype based on the ALDH2 genotyping from human subjects, we subjected participants who were heterozygous for *rs747096195* (R101G), *rs190764869* (R114W) and *rs671* (E504K) to an alcohol challenge and compared the response to those participants carrying wild type ALDH2. We considered the *rs747096195* and *rs190764869* point mutations together as these point mutations are within the same alpha helix for ALDH2, each are in extremely low frequency in humans (0.000024 for *rs747096195* and 0.0000121 for *rs190764869*)^11^, and have a similar K_m_ based on *in vitro* data. For participants given an alcohol challenge, general demographics including age, weight, and sex were collected (Supplemental Table 3). When comparing heterozygotes for *rs671* (E504K, ALDH2*1/*2) with respect to those with a wild type ALDH2 genotype, breath acetaldehyde levels peaked 9-fold 5 minutes after completing alcohol consumption (Figure 2B, 15-minute time point measurement, peak: 2.1±0.4^+^ versus 0.2±0.3 ppm, n=8/group). Further, *rs747096195* (R101G) and *rs190764869* (R114W) accumulated acetaldehyde and took longer to metabolize acetaldehyde relative to those subjects that were wild type ALDH2; supporting the recombinant enzyme studies. Additionally, skin temperature changes relative to baseline significantly increased for *rs671* at 30 minutes followed by *rs747096195* (R101G) and *rs190764869* (R114W) at 60 minutes after the start of alcohol consumption (Figure 2C). Concomitantly, heart rate changes also markedly increased in *rs671* (E504K) participants in addition to less substantial heart rate changes for those carrying *rs747096195* (R101G) and *rs190764869* (R114W) (Figure 2D).

Next, we tested additional participants that carried an intron mutation *rs4646777* (Figure 3A, G>A, intron 8, 0.18 frequency) or mixed-race heterozygotes for *rs671* (E504K). General demographics including age, weight, and sex are described (Supplemental Table 4). When subjected to an alcohol challenge, *rs4646777* had minimal changes in acetaldehyde concentrations (Figure 3B), with metabolism nearly identical to those genotyped as wild type ALDH2 (Figure 2B). However, unlike those genotyped as wild type ALDH2, noticeable facial flushing with increases in facial skin temperature occurred after an alcohol challenge for participants either carrying *rs4646777* or *rs671* (Figure 3C). The timing of facial flushing was also different between *rs4646777* or *rs671* with *rs4646777* having a more delayed facial flushing response that peaked 90 minutes after alcohol consumption. Heart rate changes after alcohol also occurred for those carrying *rs671* as opposed to *rs4646777* (Figure 3D).

**Figure 3.**
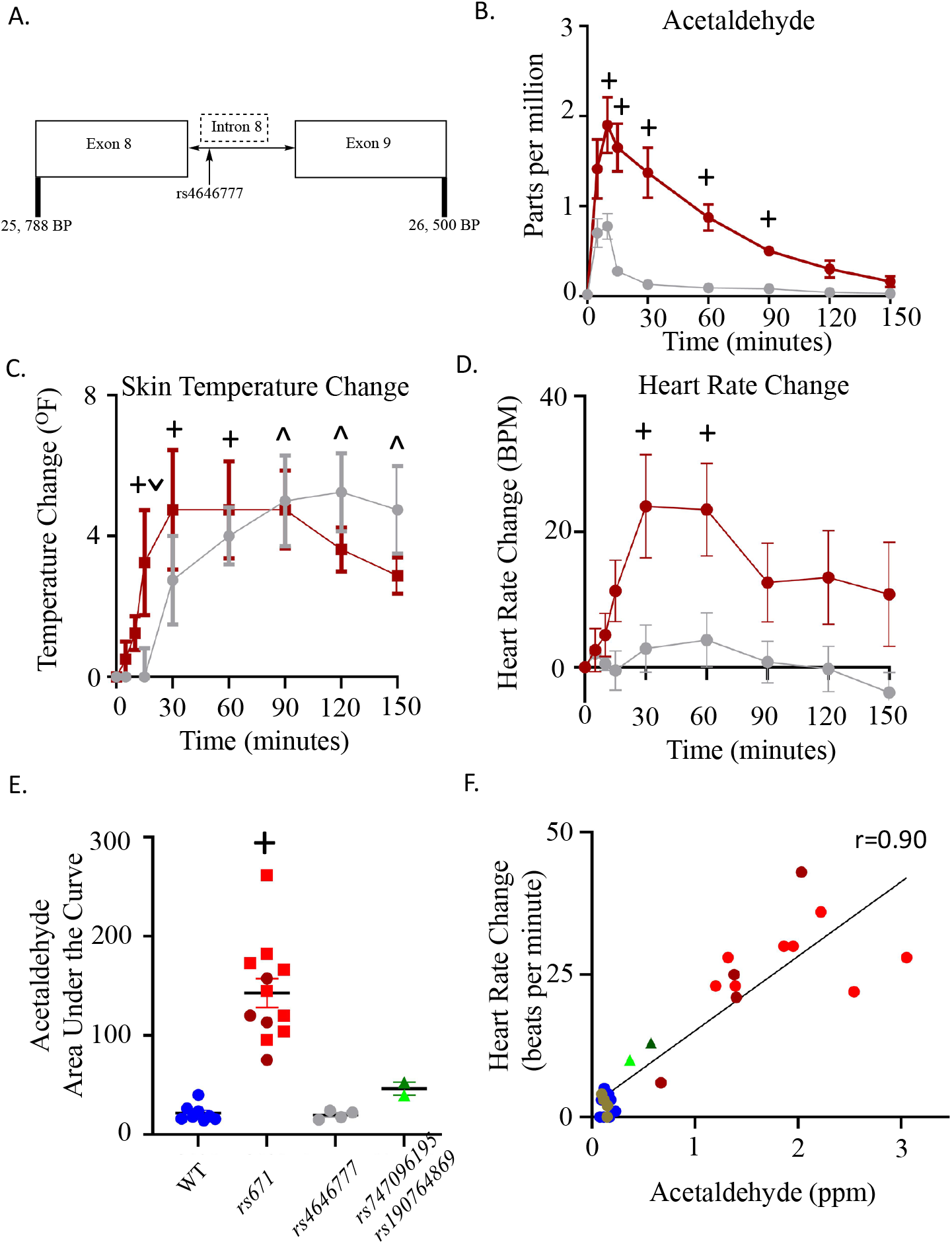
ALDH2 intron mutation *rs4646777* causes facial flushing without acetaldehyde accumulation. **A**. Intron location for *rs4646777* **B**. Acetaldehyde levels after alcohol consumption for subjects heterozygotic for *rs4646777* and subjects that were mixed-race for *rs671*. **C**. Skin temperature changes for *rs4646777* and *rs671*. **D**. Heart rate changes for *rs4646777* and *rs671*. **E**. Cumulative acetaldehyde levels after alcohol consumption for each group tested. Data is expressed as area under the curve (ppm-min). WT= wild type ALDH2. **F**. Heart rate change versus acetaldehyde levels at 30 minutes after starting alcohol consumption. *rs671* (red and dark red), *rs4646777* (grey), *rs747096195* and *rs190764869* (green), and wild type ALDH2 (blue). ^˅^P<0.01 relative to *rs4646777*, ^+^ or ^P<0.01 relative to wild type ALDH2 subjects.

Together, when comparing total acetaldehyde accumulation after an alcohol challenge for all subjects tested, acetaldehyde accumulation was 6-fold greater for heterozygotes with *rs671* and more variable versus those without ALDH2 mutations (Figure 3E, 143±14^+^ versus 22±3 ppm-minutes). Further, total acetaldehyde was minimal for people with *rs4646777* (20±2 ppm-minutes) as opposed to a two-fold accumulation for *rs747096195* and *rs190764869* (46±7 ppm-minutes) (Figure 3E). Heart rate changes 30 minutes after the start of alcohol consumption correlated with total acetaldehyde area under the curve (r=0.88, 95% confidence 0.74-0.94, R^2^=0.77, Supplemental Figure 3A) and acetaldehyde levels measured at 30 minutes after the start of alcohol consumption (r=0.90, 95% confidence 0.78-0.95, R^2^=0.80, Figure 3F). This was opposed to skin temperature changes that had less of a correlation with total acetaldehyde area under the curve (r=0.53, 95% confidence 0.19-0.76, R^2^=0.28, p<0.001, Supplemental Figure 3B) and acetaldehyde levels measured at 30 minutes after the start of alcohol consumption (r=0.55, 95% confidence 0.21-0.77, R^2^=0.30, Supplemental Figure 3C).

## Discussion

Here we identified and characterized human ALDH2 point mutations using recombinant enzyme and developed a non-invasive method to screen for inactivating ALDH2 point mutations in humans. As we find here, additional inactivating ALDH2 mutations besides *rs671* exist in humans; indicating greater than 8% of the world population are impacted by inefficient aldehyde metabolism. Importantly, people self-identifying as non-Asian also carry ALDH2 inactivating point mutations and ALDH2 inactivating point mutations are not exclusive to the Asian population. Therefore, screening for inactivating ALDH2 point mutations should be inclusive for all ethnic populations and with this approach can lead to developing precision medicine pathways that can provide more individualized recommendations regarding the health risks with aldehyde exposures. This is imperative since prior studies clearly support people carrying inactivating ALDH2 mutations, such as *rs671*, are at higher risk for cancer, cardiovascular, and neurological diseases with aldehyde exposure^4^.

Although facial flushing is commonly considered a sign for carrying an ALDH2 inactivating point mutation, our study does not support this association. Instead, monitoring heart rate changes after alcohol consumption can provide more detailed information regarding aldehyde metabolism beyond that of genetic information alone. Humans heterozygous for *rs671* are non-homogenous in their response to aldehyde metabolism; stemming from ALDH2 enzyme activity being highly dependent on the ratio between E504K and E504 monomers forming the ALDH2 tetramer^3^. Therefore, assessing heart rate changes after alcohol, as we show here, can further categorize the severity of acetaldehyde accumulation from carrying an inactive ALDH2 mutation individually. Heart rate monitoring during alcohol consumption could also open up an opportunity for people to screen for inactive ALDH2 point mutations in their home by measuring heart rate before and after alcohol consumption by palpation or with a smartwatch.

This is also the first study to characterize and phenotype the novel ALDH2 point mutations *rs747096195* (R101G) and *rs190764869* (R114W). This inefficient aldehyde metabolism is different mechanistically relative to *rs671* (E504K) as R101 and R114 are residues in a long α-helix that is in a distinctly different location to E504^17^. Recombinant enzyme activity of R101G and R114W co-expressed with wild-type ALDH2 demonstrated that the missense ALDH2 mutations have less activity (Vmax/Km) relative to wild-type ALDH2. In humans, acetaldehyde metabolism for *rs747096195* (R101G) and *rs190764869* (R114W) is reduced 2-fold rather than the 6-fold response for those carrying *rs671* (E504K); likely since the K_m_ for the ALDH2 enzyme is left unaffected by the R101G and R114W point mutations. Further, several other missense mutations exist within the exome for the ALDH2 enzyme^12^ and these ALDH2 mutations will have to be studied to understand the phenotype occurring in response to aldehyde exposure.

Additionally, *rs4646777* causes facial flushing without acetaldehyde accumulation. The pathophysiological significance of *rs4646777* is less known. One GWAS study involving a case-control study in a Norwegian population for lung cancer with 335 cases and 415 controls examining 105 SNPs identified a possible association of *rs4646777* with lung cancer which later in this study was considered a false positive and could not be validated^18^. As GWAS studies have not identified *rs4646777* as a potential cause of human pathophysiology, *rs4646777* may cause facial flushing but potentially be benign. However, as *rs4646777* has a high frequency (.18) relative to other ALDH2 mutations, it is important to further understand if there are health implications for carrying *rs4646777* besides causing facial flushing after alcohol consumption.

In the past, *rs671* carriers were considered protected from alcohol-related health problems by abstaining from alcohol use. However, even moderate alcohol consumption for *rs671* carriers increases the odds ratio for developing aerodigestive track cancer (2.61, 1.19-5.75) and esophageal cancer (3.12, 1.95-5.01)^19, 20^. This is important to recognize as societal pressures occurring globally are steering people with ALDH2 inactivating mutations towards alcohol consumption. For example, although the prevalence of *rs671* is 20% in the Republic of Korea, the work culture expects employees to attend dinners and drink alcohol. This culture likely fuels the Republic of Korea having the highest annual per capita alcohol consumption of 12.3L in the Asia Pacific region^21^. In East Asian countries, there is also a concerning trend that alcohol use disorder is steadily rising in heterozygotes who carry *rs671*^22, 23^. In the United States, social drinking and binge drinking at college is also concerning and may pressure those with ALDH2 inactivating mutations to drink alcohol^24, 25^. The COVID-19 pandemic has also seen a ∼27% increase in off-premise alcohol sales in the United States for April 2020 relative to the previous year^26^. Together, these social factors driving alcohol use need to be highlighted to raise public awareness for the importance of identifying inactivating ALDH2 point mutations that limit aldehyde metabolism when considering the carcinogenic effects of alcohol.

Our results need to be interpreted within the realm of potential limitations. As the recruitment flyers were designed to identify people that have facial flushing after they consume alcohol, it is untested whether aldehyde accumulation may occur without facial flushing. Furthermore, point mutations other than *rs671* and *rs4646777* are relatively rare in the general population, which prevented the collection of a large sample size for *rs747096195* (R101G) and *rs190764869* (R114W) that we identified. Regardless, the non-invasive assay we developed can provide individualized assessments of acetaldehyde metabolism; leading to more personalized medicine strategies by identifying those that have impaired acetaldehyde metabolism after alcohol consumption versus those that do not.

In summary, we identified novel point mutations in ALDH2 and developed a methodology to phenotype their response to alcohol. This can present an opportunity to develop precision medicine pathways and individualized recommendations for alcohol use to ultimately reduce alcohol-associated cancer risk.

## Data Availability

All data is presented within the manuscript and is freely available.

## Acknowledgements

The authors would like to thank Martina Steffen for her initial support as a clinical coordinator for this study and Kayla Anchondo-Johnson, RN for her assistance in collecting physiologic data.

## References

1. Dalleau S, Baradat M, Gueraud F and Huc L. Cell death and diseases related to oxidative stress: 4-hydroxynonenal (HNE) in the balance. Cell Death Differ. 2013;20:1615–30.

2. Sinharoy P, McAllister SL, Vasu M and Gross ER. Environmental Aldehyde Sources and the Health Implications of Exposure. Adv Exp Med Biol. 2019;1193:35–52.

3. Chen CH, Ferreira JC, Gross ER and Mochly-Rosen D. Targeting aldehyde dehydrogenase 2: new therapeutic opportunities. Physiol Rev. 2014;94:1–34.

4. Gross ER, Zambelli VO, Small BA, Ferreira JC, Chen CH and Mochly-Rosen D. A personalized medicine approach for Asian Americans with the aldehyde dehydrogenase 2*2 variant. Annu Rev Pharmacol Toxicol. 2015;55:107–27.

5. Ohsawa I, Nishimaki K, Yasuda C, Kamino K and Ohta S. Deficiency in a mitochondrial aldehyde dehydrogenase increases vulnerability to oxidative stress in PC12 cells. J Neurochem. 2003;84:1110–7.

6. Lachenmeier DW and Salaspuro M. ALDH2-deficiency as genetic epidemiologic and biochemical model for the carcinogenicity of acetaldehyde. Regul Toxicol Pharmacol. 2017;86:128–136.

7. Xiao Q, Weiner H, Johnston T and Crabb DW. The aldehyde dehydrogenase ALDH2*2 allele exhibits dominance over ALDH2*1 in transduced HeLa cells. J Clin Invest. 1995;96:2180–6.

8. Brooks PJ, Enoch MA, Goldman D, Li TK and Yokoyama A. The alcohol flushing response: an unrecognized risk factor for esophageal cancer from alcohol consumption. PLoS Med. 2009;6:e50.

9. Kamino K, Nagasaka K, Imagawa M, Yamamoto H, Yoneda H, Ueki A, Kitamura S, Namekata K, Miki T and Ohta S. Deficiency in mitochondrial aldehyde dehydrogenase increases the risk for late-onset Alzheimer’s disease in the Japanese population. Biochem Biophys Res Commun. 2000;273:192–6.

10. Seo W, Gao Y, He Y, Sun J, Xu H, Feng D, Park SH, Cho YE, Guillot A, Ren T, Wu R, Wang J, Kim SJ, Hwang S, Liangpunsakul S, Yang Y, Niu J and Gao B. ALDH2 deficiency promotes alcohol-associated liver cancer by activating oncogenic pathways via oxidized DNA-enriched extracellular vesicles. J Hepatol. 2019;71:1000–1011.

11. Karczewski KJ, Francioli LC, Tiao G, Cummings BB, Alfoldi J, Wang Q, Collins RL, Laricchia KM, Ganna A, Birnbaum DP, Gauthier LD, Brand H, Solomonson M, Watts NA, Rhodes D, Singer-Berk M, England EM, Seaby EG, Kosmicki JA, Walters RK, Tashman K, Farjoun Y, Banks E, Poterba T, Wang A, Seed C, Whiffin N, Chong JX, Samocha KE, Pierce-Hoffman E, Zappala Z, O’Donnell-Luria AH, Minikel EV, Weisburd B, Lek M, Ware JS, Vittal C, Armean IM, Bergelson L, Cibulskis K, Connolly KM, Covarrubias M, Donnelly S, Ferriera S, Gabriel S, Gentry J, Gupta N, Jeandet T, Kaplan D, Llanwarne C, Munshi R, Novod S, Petrillo N, Roazen D, Ruano-Rubio V, Saltzman A, Schleicher M, Soto J, Tibbetts K, Tolonen C, Wade G, Talkowski ME, Genome Aggregation Database C, Neale BM, Daly MJ and MacArthur DG. The mutational constraint spectrum quantified from variation in 141,456 humans. Nature. 2020;581:434–443.

12. Chen CH, Ferreira JCB, Joshi AU, Stevens MC, Li SJ, Hsu JH, Maclean R, Ferreira ND, Cervantes PR, Martinez DD, Barrientos FL, Quintanares GHR and Mochly-Rosen D. Novel and prevalent non-East Asian ALDH2 variants; Implications for global susceptibility to aldehydes’ toxicity. EBioMedicine. 2020;55:102753.

13. International Agency for Research on Cancer Monograph Working Group, Special Report: Policy A review of human carcinogens—Part E: tobacco, areca nut, alcohol, coal smoke, and salted fish. The Lancet 2009 10, 1033–1034.

14. Zambelli VO, Gross ER, Chen CH, Gutierrez VP, Cury Y and Mochly-Rosen D. Aldehyde dehydrogenase-2 regulates nociception in rodent models of acute inflammatory pain. Sci Transl Med. 2014;6:251ra118.

15. National Institutes of Health. What is a Standard Drink? NIAAA: Understanding the impact of alcohol on human health and well-being. https://www.niaaa.nih.gov/what-standard-drink, Assessed July 15, 2020.

16. Kim SW, Bae KY, Shin HY, Kim JM, Shin IS, Youn T, Kim J, Kim JK and Yoon JS. The role of acetaldehyde in human psychomotor function: a double-blind placebo-controlled crossover study. Biol Psychiatry. 2010;67:840–5.

17. Steinmetz CG, Xie P, Weiner H and Hurley TD. Structure of mitochondrial aldehyde dehydrogenase: the genetic component of ethanol aversion. Structure. 1997;5:701–11.

18. Zienolddiny S, Campa D, Lind H, Ryberg D, Skaug V, Stangeland LB, Canzian F and Haugen A. A comprehensive analysis of phase I and phase II metabolism gene polymorphisms and risk of non-small cell lung cancer in smokers. Carcinogenesis. 2008;29:1164–9.

19. Tsai ST, Wong TY, Ou CY, Fang SY, Chen KC, Hsiao JR, Huang CC, Lee WT, Lo HI, Huang JS, Wu JL, Yen CJ, Hsueh WT, Wu YH, Yang MW, Lin FC, Chang JY, Chang KY, Wu SY, Liao HC, Lin CL, Wang YH, Weng YL, Yang HC and Chang JS. The interplay between alcohol consumption, oral hygiene, ALDH2 and ADH1B in the risk of head and neck cancer. Int J Cancer. 2014;135:2424–36.

20. Yang SJ, Yokoyama A, Yokoyama T, Huang YC, Wu SY, Shao Y, Niu J, Wang J, Liu Y, Zhou XQ and Yang CX. Relationship between genetic polymorphisms of ALDH2 and ADH1B and esophageal cancer risk: a meta-analysis. World J Gastroenterol. 2010;16:4210–20.

21. Monzavi SM, Salarian AA, Khoshdel AR, Dadpour B and Afshari R. Effectiveness of a clinical protocol implemented to standardize snakebite management in Iran: initial evaluation. Wilderness Environ Med. 2015;26:115–23.

22. Yokoyama A, Muramatsu T, Ohmori T, Yokoyama T, Okuyama K, Takahashi H, Hasegawa Y, Higuchi S, Maruyama K, Shirakura K and Ishii H. Alcohol-related cancers and aldehyde dehydrogenase-2 in Japanese alcoholics. Carcinogenesis. 1998;19:1383–7.

23. Chang JS, Hsiao JR and Chen CH. ALDH2 polymorphism and alcohol-related cancers in Asians: a public health perspective. J Biomed Sci. 2017;24:19.

24. Frank E, Elon L, Naimi T and Brewer R. Alcohol consumption and alcohol counselling behaviour among US medical students: cohort study. BMJ. 2008;337:a2155.

25. Naimi TS, Brewer RD, Mokdad A, Denny C, Serdula MK and Marks JS. Binge drinking among US adults. JAMA. 2003;289:70–5.

26. The Nielson Company. Rebalancing the ‘COVID-19 effect’ on alcohol sales. Published May 7, 2020. Assessed May 22, 2020. https://www.nielsen.com/us/en/insights/article/2020/rebalancing-the-covid-19-effect-on-alcohol-sales/.

