## Supplemental Methods, Tables, Figures for "Phenotyping aldehyde metabolism in humans uncovers novel genetic mutations in aldehyde dehydrogenase 2"

for

**Supplemental Methods:**

Exclusion criteria for study: Exclusion criteria were age less than 21 years or greater than 85 years, weight greater than 225 pounds, pregnancy or breastfeeding, no prior history of alcohol consumption, medication use or current health conditions that contraindicate alcohol consumption, shortness of breath in the last month, history of severe allergic reaction to alcohol, and absence of alcohol-induced flushing after one standard drink of alcohol.

**Supplemental Tables:**

**Supplemental Table 1. Primers used to sequence an ALDH2 intron region (*rs4646777*) and ALDH2 exons.** Twelve primer sets were used, with forward and reverse primers listed.

**Supplemental Table 2. Formula used to calculate acetaldehyde using selective ion flow mass spectrometry.** The reaction ion reaction ratio and mass used for detection of acetaldehyde are listed.

**Supplemental Table 3. Human volunteer demographics.** Number of people, age, weight, sex and smoking status for volunteers genotyped as heterozygous for *rs671*, *rs747096195*, *rs190764869*, and wild type ALDH2.

**Supplemental Table 4. Additional human volunteer demographics for study.** Number of people, age, weight, sex and smoking status for volunteers genotyped as heterozygous for *rs671* with a mixed-race background and those heterozygous for *rs4646777*.

**Supplemental Figure Legends:**

**Supplemental Figure 1. Flowchart for alcohol challenge. A.** After screening and genotyping, 8 subjects (4 male and 4 female) were selected for alcohol challenge that carried a *rs671* (ALDH2*1*2) genotype. Additionally, 8 subjects wild type for ALDH2 were age and sex matched to those with an *rs671* genotype. **B.** Recruitment of subjects with additional ALDH2 mutations. After screening and genotyping, 10 participants were selected for an alcohol challenge.

**Supplemental Figure 2. Change in facial skin temperature or heart rate relative to total acetaldehyde accumulation after an alcohol challenge. A.** Change in heart rate relative to total acetaldehyde accumulated after an alcohol challenge. **B.** Change in skin temperature relative to total acetaldehyde accumulated after alcohol challenge. **C.** Change in skin temperature relative to acetaldehyde levels at 30 minutes after the start of alcohol consumption. *rs671* (red)**,** *rs4646777* (grey), *rs190764869 and rs747096195* (green), and wild type ALDH2 (blue).

Supplemental Table 1.

PCR Primer Sets for ALDH2 Exon Genotyping

| Exon | Forward Primer | Reverse Primer |
|---|---|---|
| 1 | TAGCGCCACCCGCTTCGCTTGCATCA | TGCGGGGGACTCGGGCCGGAAAACAAA |
| 2 | GGTAGTCAGTATTAGGTTGACAGCTGG | TTAACACCGTTGTCTGCAGACAAG |
| 3 | TGTGCAGCGATATGCTGATGACC | GCTGCGAGCCTATTCACCACAT |
| 4 | TTGGAGAGACCATGGCAATAGTCCAG | TTTACCTCCTAACAACGTTGCCTCCC |
| 5 | CAGAAAGACTCAGCTGGACCAGTTTG | TTGTCAGAGCCCATCTTCTTACCTGCC |
| 6 | CCAGTGTAGTTCTCTGAGGAAGCT | TAAGAGGGAGCCACTCTGGTTCACAT |
| 7 | GACCACATGTGTCCTTGGCAGA | GGAGCTCACCTTGAGCATGTCGT |
| 8 | TCTCTCGTGGTCCAGTTGCTCACT | ACAGCAACCAACGCCATTGGGCA |
| 9 | AGCAGGCATCAACCCTTACAGT | TTCTCATGCTGGCTCTAGACA |
| 10 | GGCTGCATAATTCTAAGCCTGAAGCCT | CATCAGGATGCCAGGCTGAACTGTTT |
| 11 | TTCCCCTGGAACTGTTAGAGCATGGCT | ATTCCAGGATGGTGACCACCAGATTC |
| 12 | GTCCTGGGAGTGTAACCCATAA | AACAGACCCCAATCCCCCAGCA |

Supplemental Table 2.

| Metabolite | Reagent Ion | Reaction Ratio | Mass (m/z) |
|---|---|---|---|
| Acetaldehyde | $H_3O^+$ | $3.7 \times 10^{-9}$ | 45, 63 |
| | $NO^+$ | $6.0 \times 10^{-9}$ | 43, 61 |

Supplemental Table 3.

| | Wild type | *rs671* | *rs747096195* and *rs190764869* |
|---|---|---|---|
| Number of participants | 8 | 8 | 2 |
| Age (years) | 28±2 | 29±2 | 43±18 |
| Weight (pounds) | 147±8 | 146±8 | 141±3 |
| Gender (Male/Female) | 4/4 | 4/4 | 0/2 |
| Smoker | None | None | None |

Supplemental Table 4.

| | *rs4646777* | *rs671* |
|---|---|---|
| Number of participants | 4 | 4 |
| Age (years) | 27±2 | 34±5 |
| Weight (pounds) | 138±9 | 159±22 |
| Gender (Male/Female) | 0/4 | 2/2 |
| Smoker | None | None |

Supplemental Figure 1.

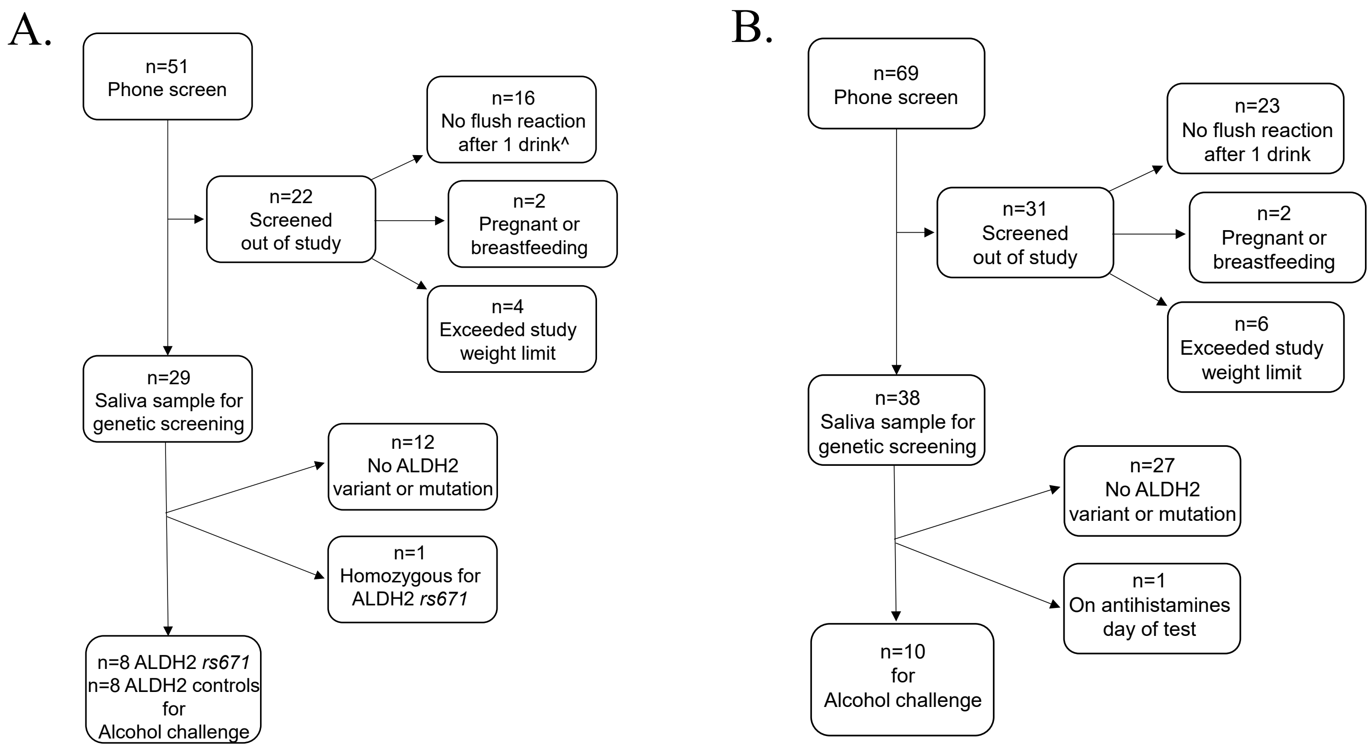

Supplemental Figure 2.

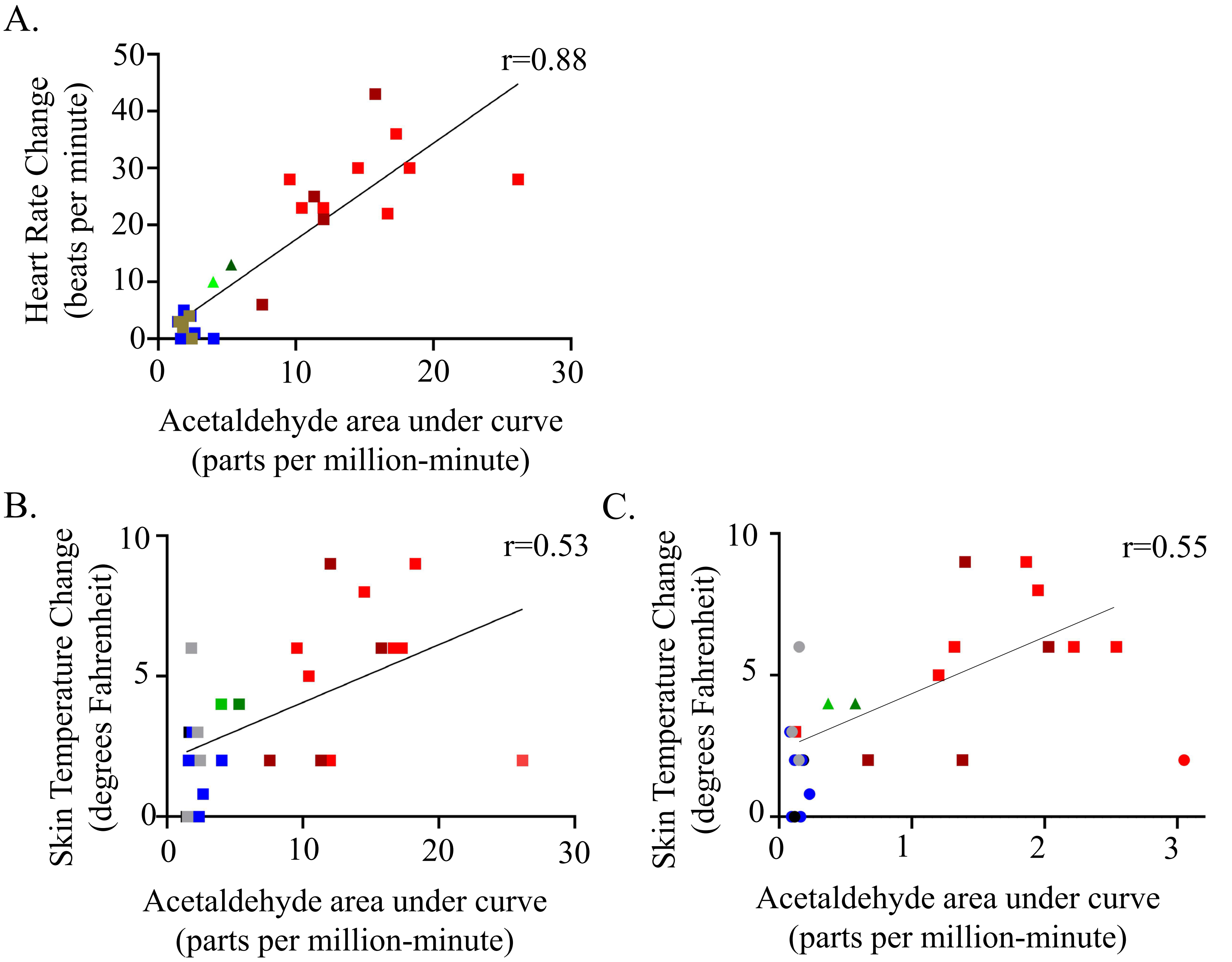
